# Remdesivir-based therapy improved recovery of patients with COVID-19 in the SARSTer multicentre, real-world study

**DOI:** 10.1101/2020.10.30.20215301

**Authors:** Robert Flisiak, Dorota Zarębska-Michaluk, Aleksandra Berkan-Kawińska, Magdalena Tudrujek-Zdunek, Magdalena Rogalska, Anna Piekarska, Dorota Kozielewicz, Krzysztof Kłos, Marta Rorat, Beata Bolewska, Anna Szymanek-Pasternak, Włodzimierz Mazur, Beata Lorenc, Regina Podlasin, Katarzyna Sikorska, Barbara Oczko-Grzesik, Cezary Iwaszkiewicz, Bartosz Szetela, Paweł Pabjan, Małgorzata Pawłowska, Krzysztof Tomasiewicz, Joanna Polańska, Jerzy Jaroszewicz

## Abstract

**Background:** Remdesivir (RDV) is the only antiviral drug registered currently for treatment of COVID-19 after a few clinical trials with controversial results. The purpose of this study was to evaluate the effectiveness and safety of RDV in patients with COVID-19 in real world settings.

**Methods:** Patients were selected from 1496 individuals included in the SARSTer national database; 122 of them received therapy with RDV and 211 were treated with lopinavir/ritonavir (LPV/r)-based therapy. The primary end-point of effectiveness was clinical improvement in the ordinal 8-point scale, which was defined as a 2-point decrease from baseline to 7, 14, 21 and 28 days of hospitalization. The secondary end-points of effectiveness included: death rate, rate of no clinical improvement within 28 days of hospitalization in the ordinal scale, rate of the need for constant oxygen therapy, duration of oxygen therapy, rate of the need for mechanical ventilation, total hospitalization time, and rate of positive RT PCR for SARS-CoV-2 after 30 days.

**Findings:** Significantly higher rates of clinical improvement, by 15% and 10% respectively, were observed after RDV treatment compared to LPV/r at days 21 and 28. The difference between regimens increased with worsening of oxygen saturation (SpO_2_) and depending on the baseline score from the ordinal scale. Statistically significant differences supporting RDV were also noted regarding the rate of no clinical improvement within 28 days of hospitalization and hospitalization duration in patients with baseline SpO_2_ ≤90%. In the logistic regression model only the administration of remdesivir was independently associated with at least a 2-point improvement in the ordinal scale between baseline and day 21.

**Interpretation:** In conclusion, data collected in this retrospective, observational, real world study supported use of remdesivir for treatment of SARS-CoV-2 infection particularly in patients with oxygen saturation ≤95%.

## Introduction

In December of 2019, a new pathogen associated with an outbreak of respiratory tract infections was discovered in Wuhan, China. It was identified as a novel coronavirus termed severe acute respiratory syndrome coronavirus (SARS-CoV-2) closely related to already known betacoronaviruses responsible for epidemics named with the acronyms SARS and MERS (Middle East respiratory syndrome). The outbreak of the disease it causes, named coronavirus disease 2019 (COVID-19), was announced a global pandemic by the World Health Organization (WHO) in March 2020. The clinical spectrum of SARS-CoV-2 infection ranges from asymptomatic or mild self-limited respiratory tract disease to severe progressive pneumonia leading to acute respiratory distress syndrome (ARDS), and death due to multi-organ failure. At the start of the epidemic, only supportive care was available, but the rapid worldwide spread of COVID-19 has raised a desperate need to invent an antiviral agent active against SARS-CoV-2. At the beginning, the search for effective therapy focused on drug repurposing and new uses for approved agents with confirmed activity against other viruses. Among them, a compound of lopinavir and ritonavir (LPV/r) was identified. Lopinavir acting as an inhibitor of a human immunodeficiency virus (HIV) protease, co-administered with ritonavir to increase its bioavailability, was demonstrated to have in vitro activity against both SARS-CoV and MERS-CoV viruses [1, 2, 3, 4, 5]. The positive impact of LPV/r on clinical outcome and reduction of the viral load in nasopharyngeal swabs were documented in the patients participating in the open-label study performed at the outbreak of SARS in 2003. Three case reports and one retrospective study described the use of LPV/r in patients with MERS, suggesting improved clinical outcome [6, 7]. Hence, due to the structural similarity of all betacoronaviruses, the relevance of LPV/r in the treatment of COVID-19 was considered. To answer the question of whether this antiviral agent works in SARS-CoV-2 infection, Cao et al. [8] performed an urgent open-label randomized clinical trial in Wuhan, the epicentre of the outbreak, to evaluate the LPV/r efficacy in patients diagnosed with COVID-19. Nevertheless, the study results were disappointing and no superiority of LPV/r therapy in terms of clinical improvement, duration of hospitalization, or period of viral RNA detectability as compared to standard of care (SoC), was documented. The only difference of statistical significance was observed in the median time to clinical improvement calculated after exclusion of patients with early death; however, the authors themselves called this difference “modest” [8]. No positive impact of LPV/r therapy was documented in critically ill patients with SARS-CoV-2 related pneumonia treated in the intensive care unit and also in patients with mild to moderate form of COVID-19 compared to SoC or adjuvant therapy [9, 10, 11].

Since the process of discovery, testing and registration of a new antiviral drug is long-lasting and cost-intensive with an uncertain chance of success, attention has been paid to investigational drugs with potential activity against SARS-CoV-2. The most promising among them is remdesivir (RDV), a prodrug of an adenosine nucleoside analogue which terminates viral RNA synthesis by inhibition of RNA-dependent RNA polymerase (RdRP), with established dosing and safety profile. The primary clinical indication of RDV was Ebola virus disease (EVD). However, despite the encouraging results of the in vivo efficacy evaluation in an animal model, the randomized phase III clinical trial did not clearly confirm the relevance of RDV in humans and then this investigational agent was shelved [12]. There was a renewed interest in RDV concerning SARS-CoV and MERS-CoV infections and confirmed in vitro and animal models activity against the family of coronaviruses has raised hope for effective application in the treatment of COVID-19 [13]. Since the inhibitory effect on the recently emerged novel coronavirus was demonstrated in vitro, clinical trials and compassionate use programs have started, and the WHO announced the launch of a trial that would include one group of patients treated with remdesivir [14, 15, 16]. Finally, based on findings from phase III clinical trials ACTT-1 and SIMPLE-severe, on 1st May 2020 RDV received emergency use authorization (EUA) for the treatment of COVID-19 issued by the Food and Drug Administration (FDA) [17, 18, 19]. Subsequent conditional licences in different countries worldwide have made remdesivir the first approved antiviral agent for the treatment of COVID-19.

The purpose of the study is evaluation of the effectiveness and safety of RDV administered to patients with COVID-19 in real-world settings. Lopinavir/ritonavir based regimen was used as a comparator instead of undefined and inperfect concept of "standard of care”, applied usually in COVID-19 studies.

## Material and Methods

The study population consisted of patients selected from 1496 individuals included in the SARSTer national database. This ongoing project, supported by the Polish Association of Epidemiologists and Infectiologists, is a national real-world experience study assessing treatment in patients with COVID-19. Patients whose data were collected in the SARSTer database were treated in 30 Polish centres, between 1 March and 31 August 2020. The decision about the treatment regimen was taken entirely by the treating physician with respect to current knowledge and recommendations of the Polish Association of Epidemiologists and Infectiologists [20, 21]. The current analysis included 333 adult patients who received regimens based on the antiviral drugs remdesivir or lopinavir/ritonavir administered for therapy of COVID-19. RDV was administered intravenously once a day with a loading dose of 200 mg and later a maintenance dose of 100 mg for 5–10 days. LPV/r was administered orally in a dose of 400/100 mg every 12 hours for up to 28 days. LPV/r was used mostly at the beginning of the pandemic and due to documented later lack of effectiveness is considered as a comparator for RDV in this study.

Data were entered retrospectively and submitted online by a web-based platform operated by “Tiba” sp. z o.o. Parameters collected at baseline included age, gender, body mass index (BMI), comorbidities and concomitant medications, clinical status at admission, additional medication dedicated to COVID-19, and adverse events. Baseline clinical status at admission to hospital was classified as asymptomatic, symptomatic stable with oxygen saturation (SpO_2_) >95%, symptomatic unstable with SpO_2_ 91-95%, symptomatic unstable, SpO_2_ ≤90% or acute respiratory distress syndrome (ARDS).

The primary end-point of treatment effectiveness was clinical improvement in the ordinal scale based on WHO recommendations modified to fit the specificity of the national health care system. Improvement was defined as a 2-point decrease from baseline to 7, 14, 21 and 28 days of hospitalization. The ordinal scale was scored as follows: 1. unhospitalized, no activity restrictions; 2. unhospitalized, no activity restrictions and/or requiring oxygen supplementation at home; 3. hospitalized, does not require oxygen supplementation and does not require medical care; 4. hospitalized, requiring no oxygen supplementation, but requiring medical care; 5. hospitalized, requiring normal oxygen supplementation; 6. hospitalized, on non-invasive ventilation with high-flow oxygen equipment; 7. hospitalized, for invasive mechanical ventilation or ECMO; 8. death.

The secondary end-points of effectiveness included: death rate, rate of no clinical improvement within 28 days of hospitalization in the ordinal scale, rate of the need for constant oxygen therapy, duration of oxygen therapy, rate of the need for mechanical ventilation, total hospitalization time, and rate of positive RT PCR for SARS-CoV-2 after 30 days of hospitalization.

### Statistical analysis

The results are expressed as mean ± standard deviation (SD) or n (%). P values of <0.05 were considered to be statistically significant. The significance of difference was calculated by chi-square or Fisher’s exact test for nominal variables and by Mann-Whitney U and Kruskal-Wallis ANOVA for continuous variables. Univariate comparisons were calculated by GraphPad Prism 5.1 (GraphPad Software, Inc., La Jolla, CA). Forward stepwise logistic regression models with the Bayesian information criterion (BIC) as a model selection criterion were performed with ≥2-point decrease in the ordinal scale between baseline and day 21 of hospitalization as the dependant variable. Among independent variables tested were age, sex, BMI, diabetes, coronary artery disease, baseline classification and baseline score in ordinal scale, as well as therapy with RDV, LPV/r, tocilizumab, dexamethasone, chloroquine/hydroxychloroquine, heparin, dexamethasone, convalescent plasma and azithromycin. Logistic regression models were calculated by use of R and MATLAB (MathWorks, MA, USA).

## Results

Among 333 patients included in the study 122 received therapy with RDV and 211 were treated with LPV/r. Groups were balanced regarding gender, age and BMI, but in both arms, there was a predominance of males (Table 1). Patients treated with RDV more frequently demonstrated a symptomatic unstable course of the disease with SpO_2_ ≤95% at admission to the hospital (69%) compared to those receiving LPV/r (57%), but the difference was not statistically significant. Prevalence of accompanying diseases was higher among patients treated with RDV, but the difference was significant only regarding ischaemic heart disease (Table 1). During the therapy with RDV additional medication more frequently included dexamethasone, convalescents plasma, and low molecular weight heparin, whereas LPV/r was more frequently administered together with chloroquine and azithromycin (Table 1).

**Table 1.**
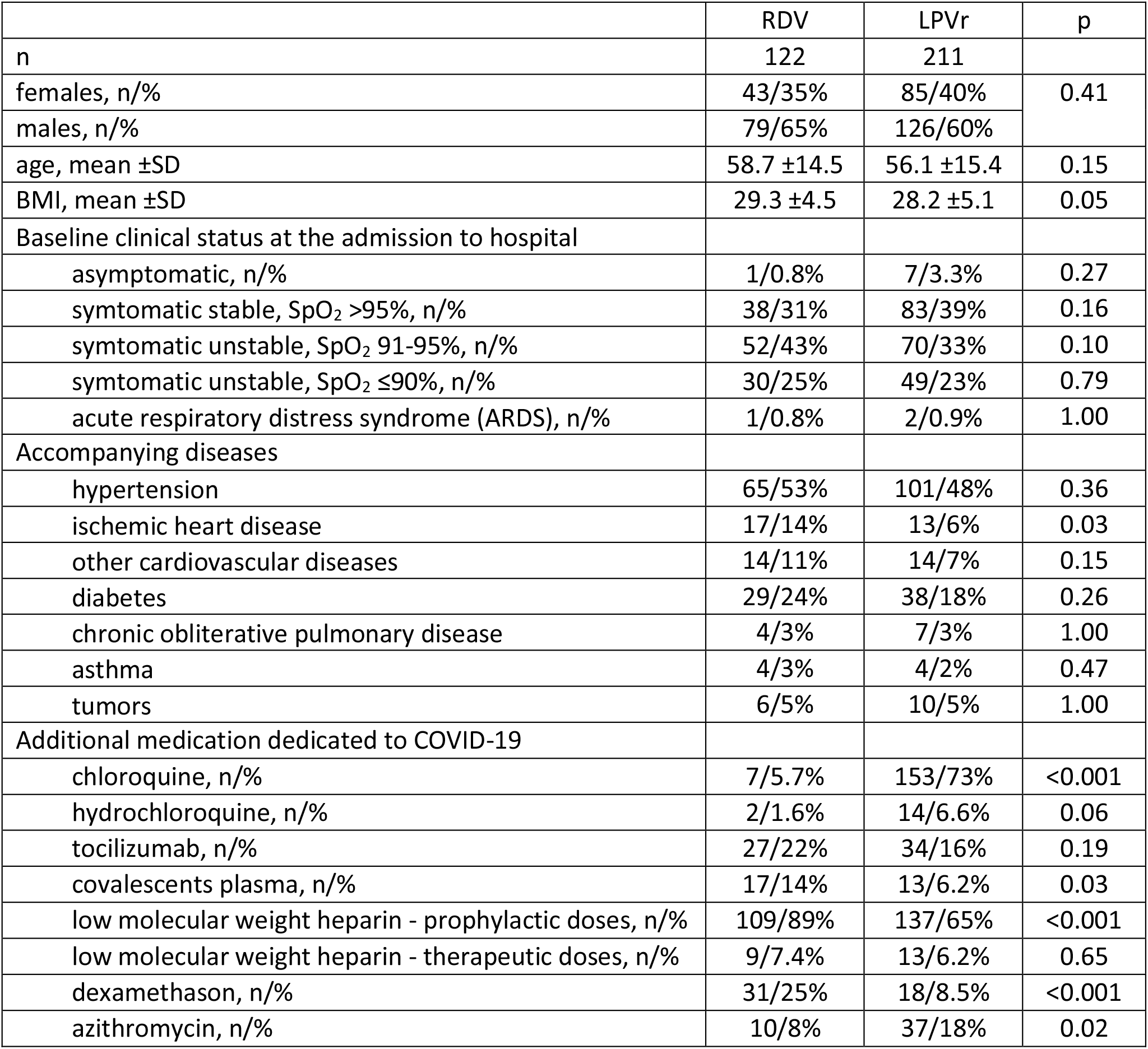
Characteristics of patients included in the study

As shown in Figure 1, proportions of categories of ordinal scale were balanced at the baseline between the two treatment groups. The rate of patients discharged from the hospital was similar on days 7 and 14, but became higher in patients treated with RDV on days 21 and 28 (Figure 1). Differences in the score in the ordinal scale between particular time points and baseline at admission to the hospital are shown in Figure 2. Clinical improvement measured using an ordinal scale demonstrated significantly higher rates after RDV treatment compared to LPV/r at weeks 21 and 28 and the difference was 15% and 10% respectively (Table 2). Additional analysis of clinical improvement using the ordinal scale was carried out depending on the baseline oxygen saturation at admission to hospital. Difference between regimens increases with worsening of oxygen saturation during the analysis on days 14, 21 and 28 (Table 2). A similar tendency was demonstrated depending on the baseline score from the ordinal scale. On the other hand, the rate of no improvement within 28 days was significantly higher after treatment with LPV/r (Table 2). This tendency was also demonstrated in analysis carried out with regard to baseline oxygen saturation or baseline ordinal score, and differences between two regimens increased with either decline of oxygen saturation or decrease of baseline ordinal score (Table 2). As shown in Table 2 the rate of no clinical improvement within 28 days of hospitalization and hospitalization duration was significantly lower in patients treated with RDV if they had baseline SpO2 ≤90%..

**Table 2.**
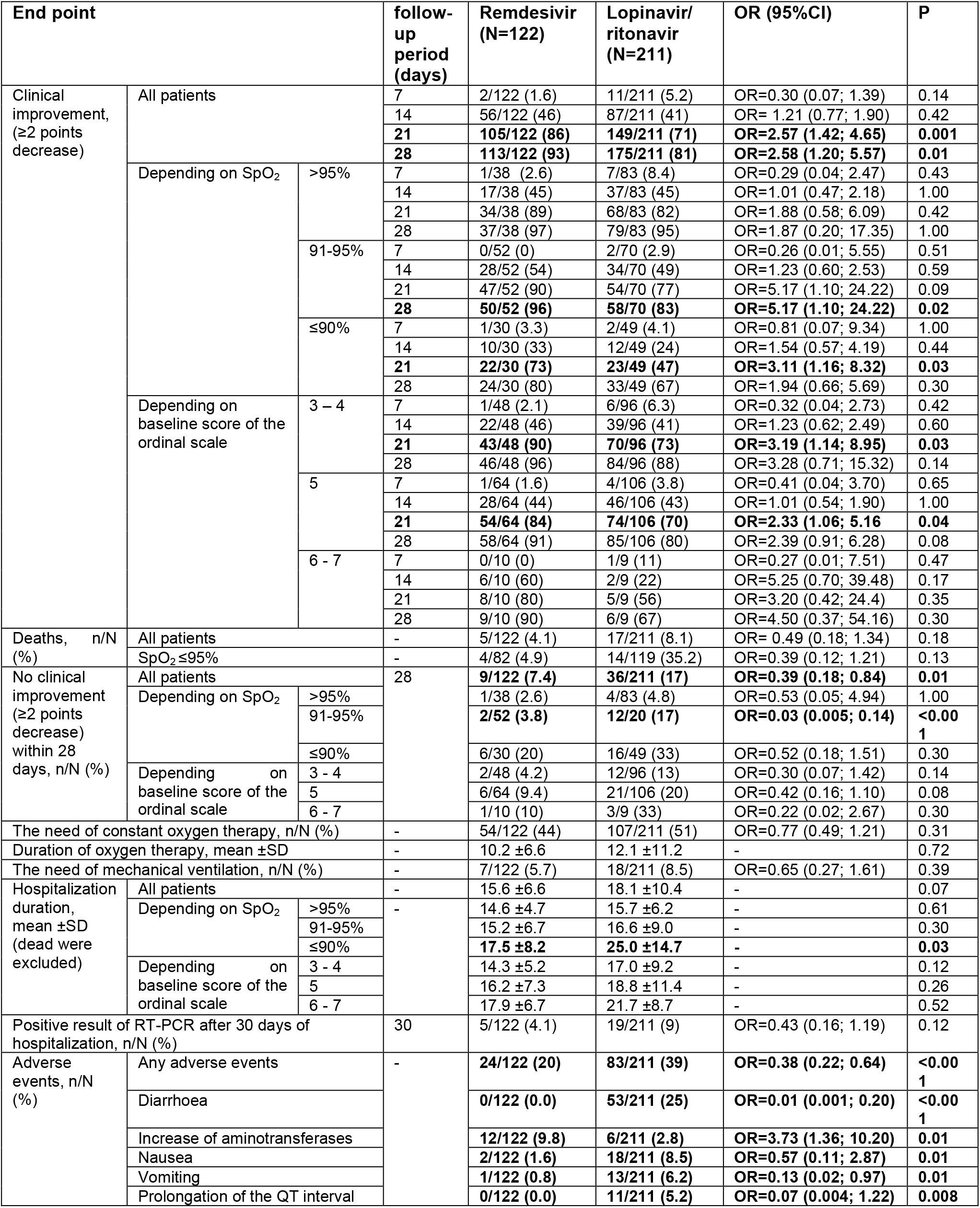
Effectiveness and safety of treatment with remdesivir compared to lopinavir/ritonavir.

**Figure 1.**
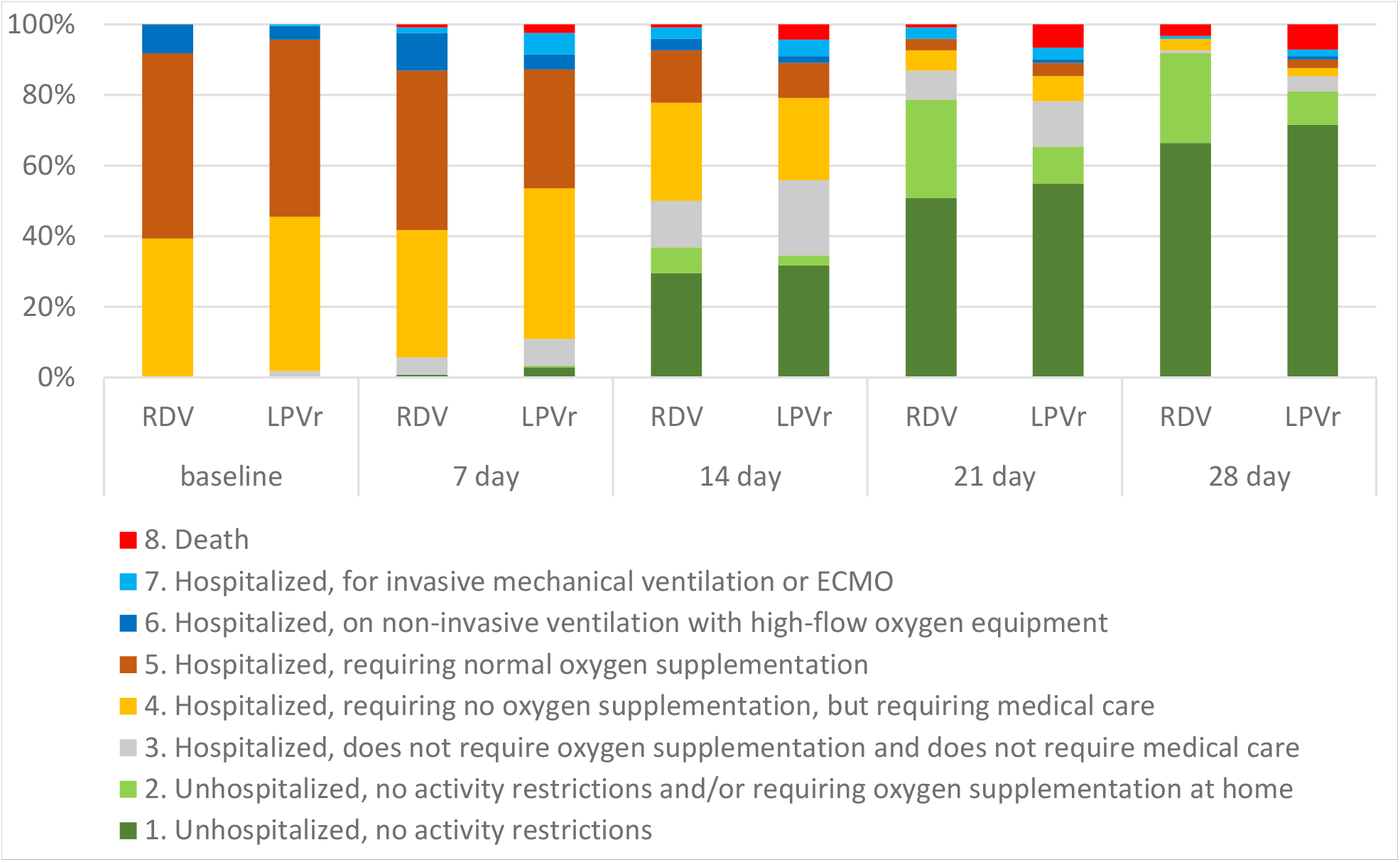
Categories of ordinal scale established in following time-points in patients treated with either remdesivir or ledipasvir/ritonavir based regimens.

**Figure 2.**
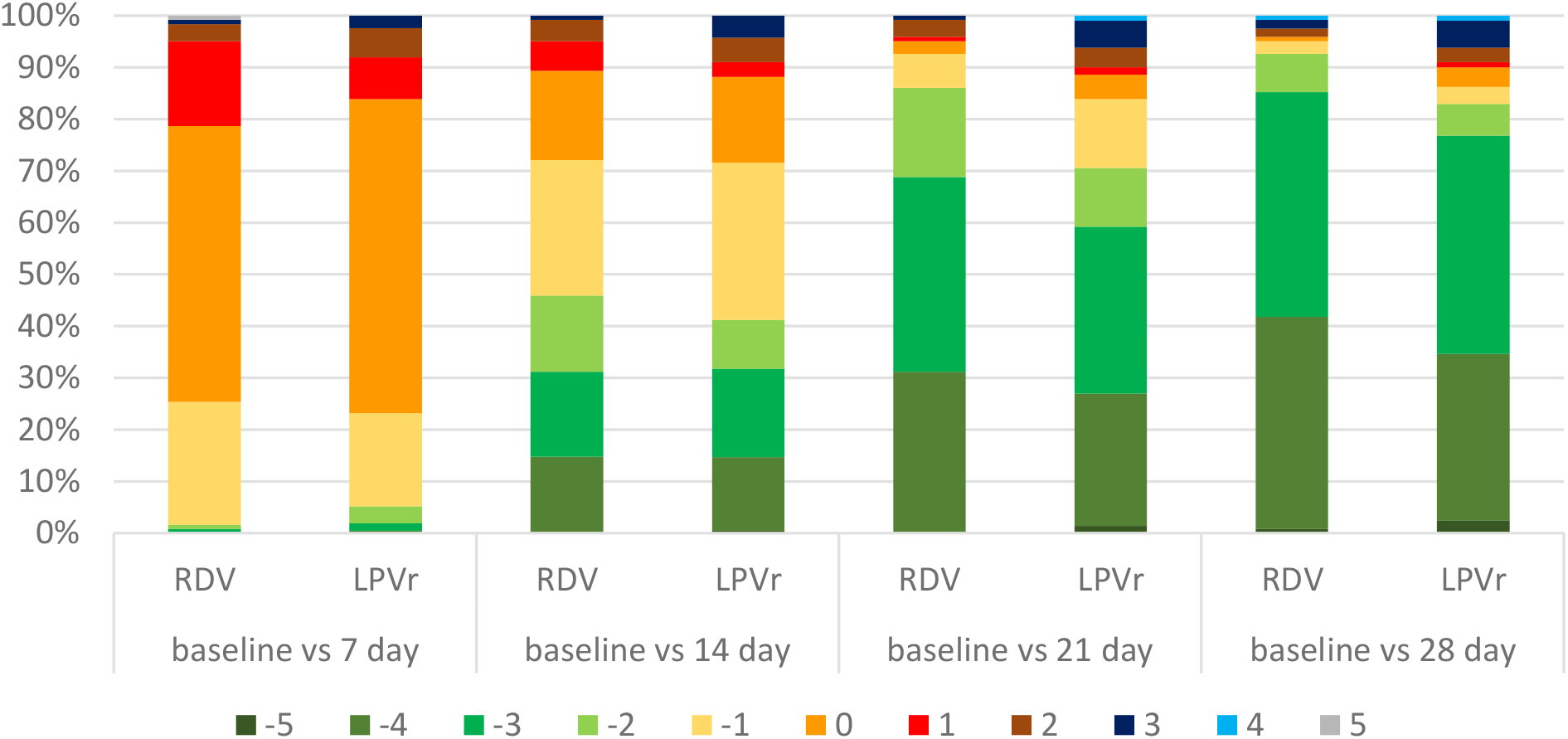
Changes in the scoring of the ordinal scale at subsequent observation time points compared to baseline.

In the logistic regression model in patients receiving RDV or LPV/r only the administration of remdesivir was independently associated with at least a 2-point improvement in the ordinal scale between baseline and day 21, while older age and use of tocilizumab were negative predictors of response (Table 3). Interestingly, in patients with older age tocilizumab on the contrary improved likelihood of response.

**Table 3.**
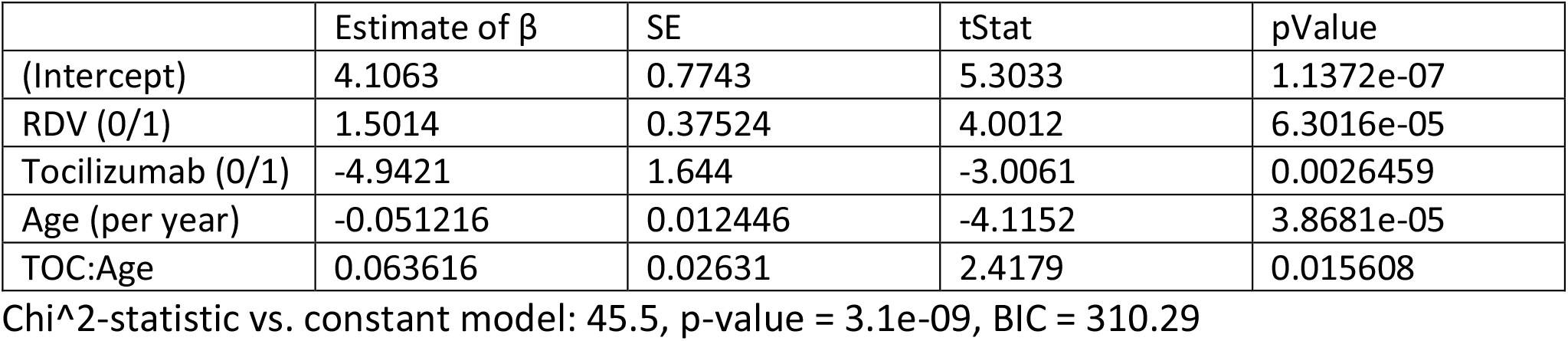
Results of the logistic regression model for improvement according to the ordinal scale, defined as ≥2 point decrease between baseline and day 21 of hospitalization in patients receiving RDV.

Patients treated with LPV/r experienced significantly more adverse events (39%) than those treated with RDV (20%). The most frequent among those receiving LPV/r were diarrhoea (25%), nausea (8.5%), vomiting (6.2%) and prolongation of the QT interval (5.2%). Patients treated with RDV most frequently experienced elevation of aminotransferases (9.8%), and other adverse events occurred sporadically (Table 2).

## Discussion

Since 1^st^ May 2020, the FDA’s emergency use authorization has allowed for remdesivir to be distributed and administered intravenously to treat COVID-19 in patients with severe disease. It has become possible due to the promising results of phase III clinical trials. Hence, the antiviral agent which failed to live up to expectation in the treatment of Ebola virus disease has become a hope for those infected with SARS-CoV-2. Conditional approvals issued in other countries and regions worldwide in the wake of the decision of the United States federal agency has enabled the treatment of RDV in patients with low blood oxygen levels or needing oxygen therapy or more intensive breathing support. However, giving access to a new potential therapy, the need for further research to evaluate the safety and effectiveness of the RDV was highlighted.

Since June 2020 remdesivir has also been available in Poland. Therefore we aimed to assess the efficacy and tolerability of RDV in real-world experience. LPV/r based regimen was used as a comparator due to its antiviral mode of action because both agents act by inhibiting viral proteins. We used this idea instead of undefined and inperfect concept of "standard of care”, applied usually in COVID-19 studies, which can be recognized in the different way depending on treating center and period of pandemic. As the lack of benefit of LPV/r treatment compared to SoC was documented by Cao et al. [8] in patients with COVID-19-related pneumonia, in the current analysis we assumed the efficacy of LPV/r at the level close to placebo. The majority of patients receiving RDV presented symptomatic unstable status with SpO_2_ ≤ 95% at admission to the hospital (68%). Using the ordinal scale widely applied in COVID-19 trials, we confirmed that RDV treatment was associated with significantly greater clinical improvement, defined as a two-point decrease in disease severity, by day 21 (86% vs. 71%, p=0.001) and 28 (93% vs. 83%, p=0.01) compared to the LPV/r arm. A significant difference at day 21 was observed regardless of the baseline score at admission to the hospital. The detailed analysis carried out considering the baseline oxygen saturation demonstrated a statistically significant difference among unstable patients, by day 21 for individuals with SpO_2_ ≤ 90%, and by day 28 for those with baseline SpO_2_ 91-95%. The possible reason for the lack of difference by day 7 and 14 could be an effect of national regulation used until 31 August 2020, which in practice ordered hospitalization of the majority of patients for at least 14 days irrespective of treatment and clinical improvement.

Hence, our findings supported results from clinical trials, including those based on which the decision of EUA was made [17, 18, 19]. Patients with COVID-19 without reduced oxygen levels included in the SIMPLE-moderate phase III trial treated with RDV for 5 days were significantly more likely to have clinical improvement as compared to those receiving SoC alone, whereas for those randomized to the 10-day regimen a significant difference was not documented [22]. Similar results were obtained for patients with the severe form of COVID-19 defined as oxygen saturation <95% while breathing ambient air or receiving oxygen support, treated with RDV in SIMPLE-severe phase III trial. Clinical improvement was demonstrated in 65% and 54% of patients receiving RDV for 5 and 10 days, respectively [18]. In both trials the assessment was performed at a time point different from ours, at day 11 for moderate and 14 for severe disease [18, 19, 22]. Notably, the comparison concerning patients with severe COVID-19 was not randomized and performed not within one study but between individuals on RDV treatment included in the SIMPLE-severe phase III clinical trial and concurrent retrospective real-world cohort study on SoC only [19]. Moreover, not all patients treated with RDV were included in the comparative analysis. After exclusion of Italian patients on RDV due to lack of comparative individuals on SoC from the real world experience (RWE) study, 74% of those treated with RDV fulfilled criteria for clinical improvement as compared to 59% among the SoC cohort at day 14 (p<0.001).

This imperfection was not shared by another phase III study ACTT-1 designed as a double-blind, randomized and placebo-controlled trial. The preliminary results of this study confirmed RDV superiority relative to placebo (normal saline solution) in shortening the time to recovery, 11 vs. 15 days. Patients were assessed daily using an ordinal scale and the primary outcome measure was the first day on which the patient reached one of the three lowest levels in the scale meaning “not hospitalized with or without limitation of activities” and “hospitalized not requiring supplemental oxygen and ongoing medical care”. However, the trial is not completed and final data are still expected [17]. Also, data from the compassionate-use programme confirmed the benefit of RDV treatment in patients with severe COVID-19, including those on invasive ventilation, reporting a 68% rate of clinical recovery [23]. Unlike the above-mentioned studies, no positive impact of RDV relative to placebo was documented in patients with the severe form of COVID-19 with baseline oxygen saturation < 95% included in the randomized, double-blind multicentre phase III clinical trial conducted in Wuhan [14]. However, it should be noted that enrolment was prematurely terminated due to control of the epidemic in Wuhan, and hence the study was underpowered.

The current analysis demonstrated benefit from RDV treatment as compared to LPV/r in terms of secondary end-points. Among them, a statistically significant difference was found for rate of no improvement within 28 days in the ordinal scale, whereas for the remaining parameters, including death rate, the need for constant oxygen therapy and its duration, the need for mechanical ventilation, rate of positive RT PCR for SARS-CoV-2 after 30 days, and hospitalization length, the positive impact of RDV without statistical significance was documented. Interestingly, most striking and significant effect of RDV on the length of hospitalization was observed in patients with baseline SpO2 who needed on average 7 days less of hospital stay. Our findings on mortality are in line with results achieved in the clinical trials SIMPLE and ACTT-1 and an observational study conducted among patients with cancer [17, 19, 22, 23].

Unfortunately, detailed comparison regarding remaining outcomes is not possible due to different end-points and time of assessment [17, 19, 22].

The tolerability profile of remdesivir in the current analysis was in accordance with clinical trials with the most frequent transient aminotransferases elevation reported in nearly 10% of patients. LPV/r was frequently responsible for transient diarrhoea, well known from experience in HIV infected patients, which did not affect significantly clinical status of patients and duration of hospitalization.

We are aware of the limitations of our study. Among them, the impact of other therapeutic agents dedicated to COVID-19 should be pointed out. Baseline characteristics demonstrated the different distribution of patients with respect to concomitant drugs with a significantly higher rate of dexamethasone, convalescent plasma and low molecular weight heparin, and a lower rate of chloroquine and azithromycin among patients treated with RDV. To eliminate this imbalance as a confounding factor we performed analysis using logistic regression, demonstrating that in patients receiving RDV or LPV/r only the administration of remdesivir was independently associated with at least a 2-point improvement in the ordinal scale at day 21. Importantly, in this model we could not prove an additional independent beneficial effect of dexamethasone or convalescent plasma on this endpoint, while the use of tocilizumab was only beneficial in older patients.

Other limitations of the current analysis are related to the RWE nature of the research, including its observational nature and retrospective electronic data capture with possible data entry errors. Unfortunately, we were not able to increase the number of patients in the study because of changed national regulations, which allowed the release of a patient from the hospital without RT PCR negativisation, which could significantly affect ordinal scale interpretation. However, the major strength of the study is collection of data from a real-world, heterogeneous population, thus being representative for routine practice, and a clearly defined comparator, unlike in some other studies, which usually used an undefined SoC. Interestingly, we observed that addition of tocilizumab can improve the likelihood of improvement, which supports our previous experience with this drug, but possible use of combined therapy of remdesivir and tocilizumab needs further study [24].

In conclusion, data collected in this retrospective, observational, real-world study with antivirals as a leading therapy in two competing arms supported the use of remdesivir compared to lopinavir/ritonavir for treatment of SARS-CoV-2 infection, particularly in patients with oxygen saturation ≤95%.

## Data Availability

All data referred can be provided by authors

## Ethics Committee

The study was retrospective, non-interventional, based on data collected in national database, so it does not require approval of Ethics Committee.

## Authors contribution

All authors had full access to all the data in the study and take responsibility for its integrity and the accuracy of the analysis. RF and DZM were responsible for the study concept, design, acquisition of data, verification of the underlying data and drafting the manuscript. JJ and JP were responsible for statistical analysis. All other authors were responsible for the acquisition of data and were involved in the final mansucript preparation.

